# Screening for Asymptomatic Carotid Artery Stenosis: A Draft Update of the Systematic Review Adapted from the USPSTF Review for the Japan Preventive Services Task Force

**DOI:** 10.1101/2025.06.26.25330063

**Authors:** Yusuke Saishoji, Tomoharu Suzuki, Saori Nonaka, Shunsuke Suya, Takashi Kitagawa

**Affiliations:** Department of General Medicine, Hakujyuji Hospital, Fukuoka, Japan; Department of Hospital Medicine, Urasoe General Hospital, Okinawa, Japan; Department of Radiation Health Management, Fukushima Medical University School of Medicine, Fukushima, Japan; Department of General Medicine, Taito Hospital, Japan Association for Development of Community Medicine, Tokyo, Japan; Department of General Internal Medicine, Hashimoto Municipal Hospital, Wakayama, Japan; Department of Physical Therapy, School of Health Sciences, Shinshu University, Nagano, Japan

**Author notes:** **Corresponding author**, Yusuke Saishoji, MD, MHA, Department of General Medicine, Hakujyuji Hospital, 4-3-1 Ishimaru, Nishi-ku, Fukuoka-shi, Fukuoka, 819-8511, Japan.

**Keywords:** Asymptomatic Carotid Artery Stenosis, Screening, Stroke Prevention, Systematic Review, Revascularization

## Abstract

**Background:** Carotid artery stenosis is a notable risk factor for ischemic stroke. In 2021, the U.S. Preventive Services Task Force (USPSTF) published an updated evidence review on screening for asymptomatic carotid artery stenosis (ACAS). At the request of the Japan Preventive Services Task Force, we conducted a systematic review to adapt and update the USPSTF review by incorporating recent international evidence and Japanese-language literature.

**Methods:** Following the USPSTF analytic framework, we evaluated the evidence on screening effectiveness, harms of screening or confirmatory testing, the incremental benefit of revascularization beyond current medical therapy, and harms of surgical interventions in asymptomatic individuals. The International Medical Information Center conducted literature searches in PubMed, the Cochrane Library, and Ichushi-Web. Searches were limited to English and Japanese studies. Two reviewers independently performed study selection, data extraction, and risk of bias assessment, with disagreements resolved by consensus. Newly identified studies were qualitatively synthesized with the 2021 USPSTF findings.

**Results:** No eligible studies directly assessing the benefits and harms of screening for ACAS were identified. One RCT evaluated the benefits of revascularization, and harms were assessed in that RCT and five observational studies. The RCT (SPACE-2; n=513), which investigated the incremental benefit of revascularization, was terminated prematurely and had substantial methodological limitations. In five registry- or claims-based observational studies, the 30-day incidence of stroke or death following revascularization was 2.5% to 2.8%. Perioperative stroke, death, and myocardial infarction occurred in 0.9% to 2.3%, 0.3% to 0.9%, and 0.3% to 0.9% of patients, respectively, consistent with the 2021 USPSTF review.

**Conclusions:** There is no direct evidence evaluating the effectiveness and harms of screening for asymptomatic carotid artery stenosis. Evidence on the benefits and harms of adding revascularization to optimal medical therapy was limited by early trial termination and methodological concerns, reducing the internal validity of the available data.

## Background

Carotid artery stenosis is a recognized cause of ischemic stroke ^1, 2^. In Japan, stroke persists as a leading cause of mortality, highlighting the critical importance of effective prevention strategies for public health. The management of asymptomatic carotid artery stenosis (ACAS) presents a clinical dilemma regarding the appropriateness of preventive intervention.

In 2021, the U.S. Preventive Services Task Force (USPSTF) issued a Grade D recommendation against screening for ACAS in the general asymptomatic adult population ^3^. This recommendation was based on the low prevalence of ACAS, the potential harms from false-positive results leading to unnecessary procedures, and the uncertain net benefit of invasive interventions when added to modern medical therapy. However, the applicability of these findings to the Japanese population is not well established. Differences in population demographics, such as advanced aging, alongside variations in stroke incidence, healthcare infrastructure, and screening practices, may alter the benefits and harms of ACAS screening in a Japanese context.

Therefore, this review aimed to provide an up-to-date and objective assessment of the current evidence on the screening and management of asymptomatic carotid artery stenosis in Japanese primary care, with a particular focus on internal validity.

## Methods

This study was conducted in accordance with the Preferred Reporting Items for Systematic Review and Meta-Analysis (PRISMA) guidelines ^4^.

### Scope of Review

This review was guided by the analytic framework depicted in Figure 1. The framework and its Key Questions (KQs) were adapted from the 2021 USPSTF evidence review ^5^ and subsequently revised to align with the Japanese clinical context.

**Figure 1.**
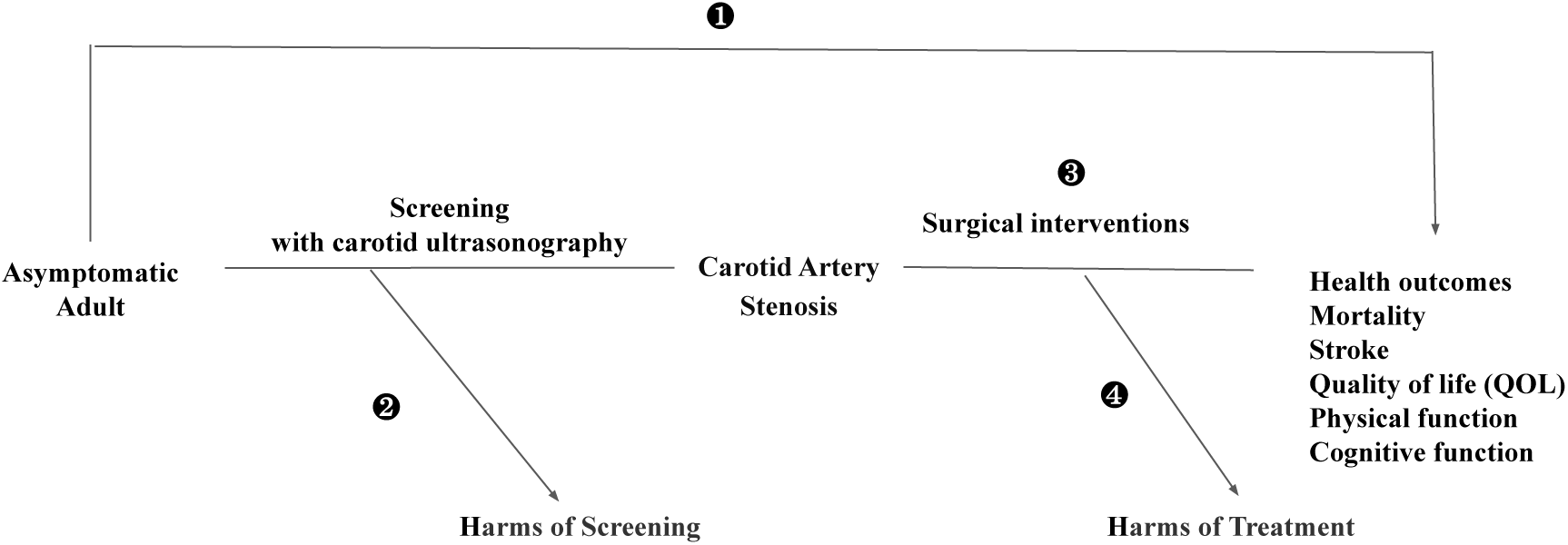
Analytic Framework: Screening for Asymptomatic Carotid Artery Stenosis.

### Data Sources and Searches

Literature searches were conducted by the International Medical Information Center. PubMed and the Cochrane Library were searched for studies published in English or Japanese between February 19, 2020, and November 26, 2024. The Ichushi-Web (Japan Medical Abstracts Society) database was searched without restriction on publication date. Detailed search strategies are provided in Table 1.

**Table 1.**
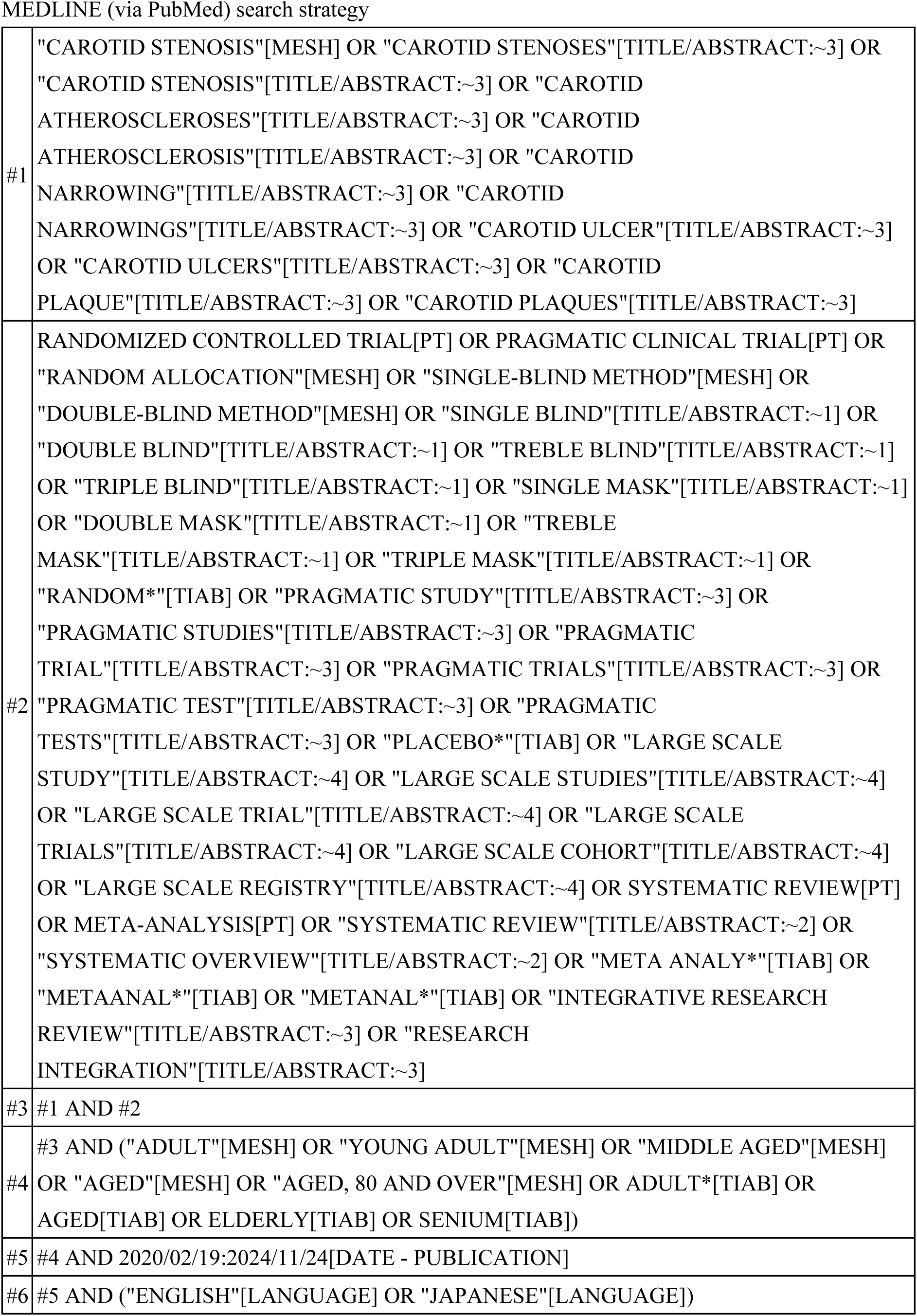
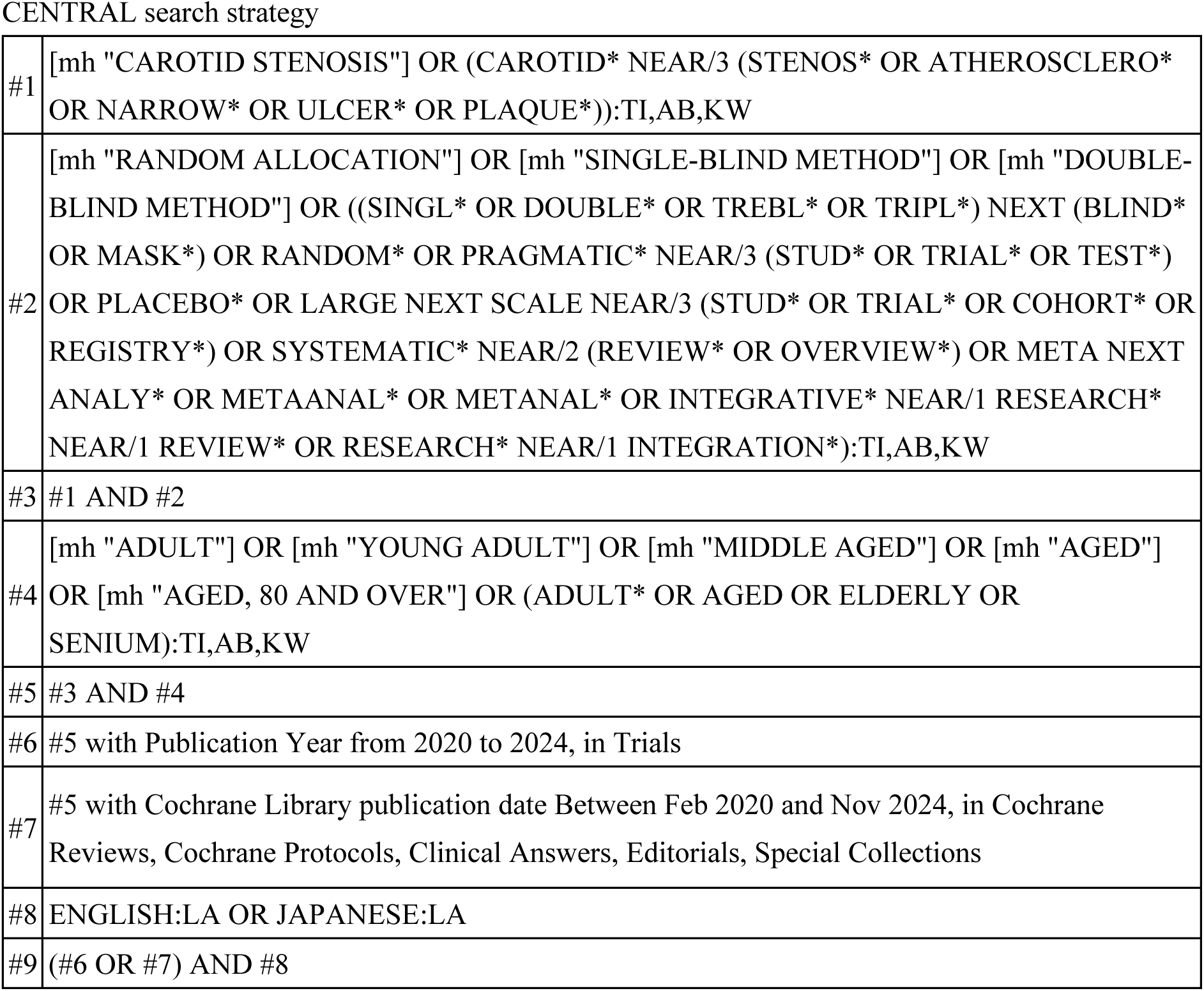
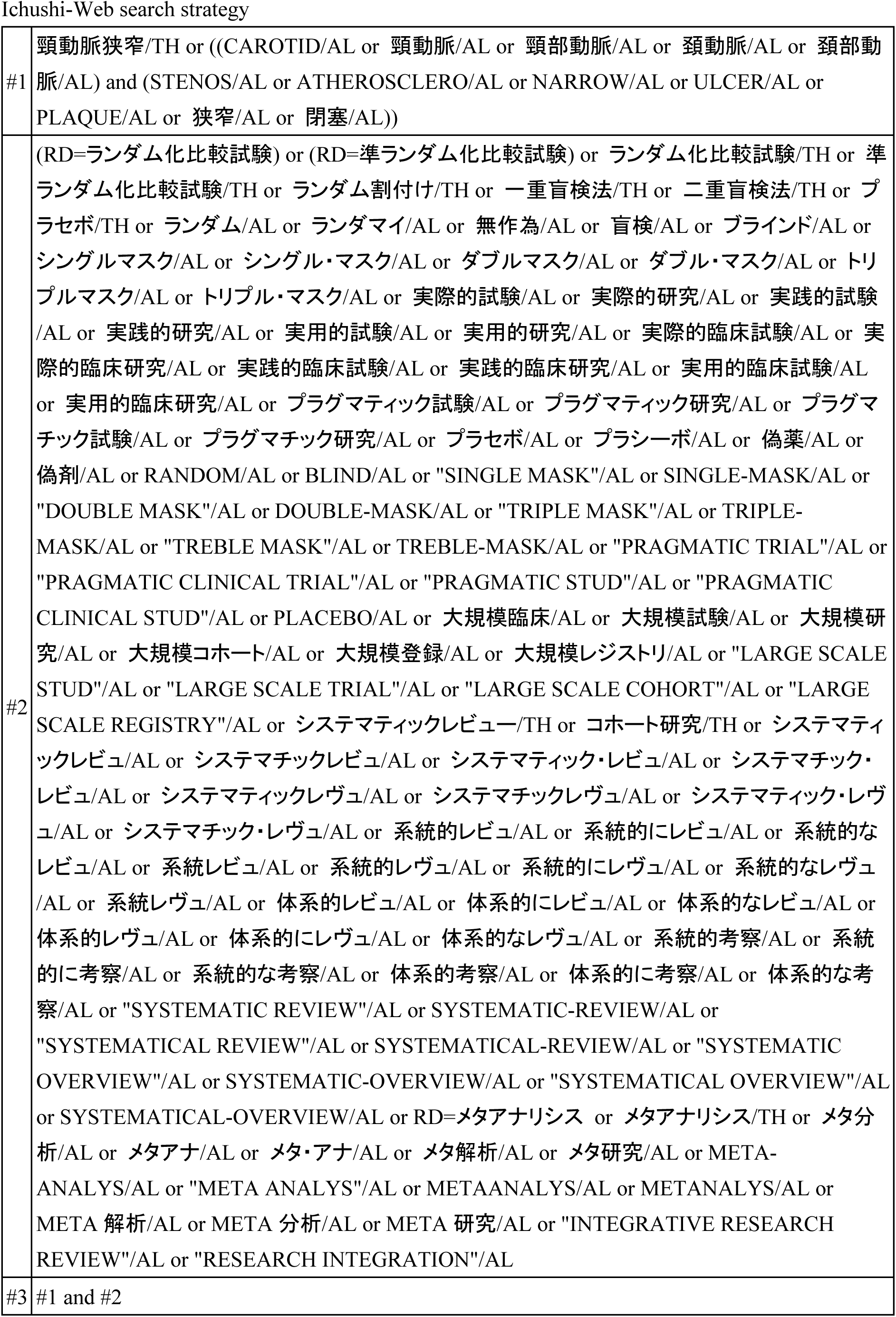
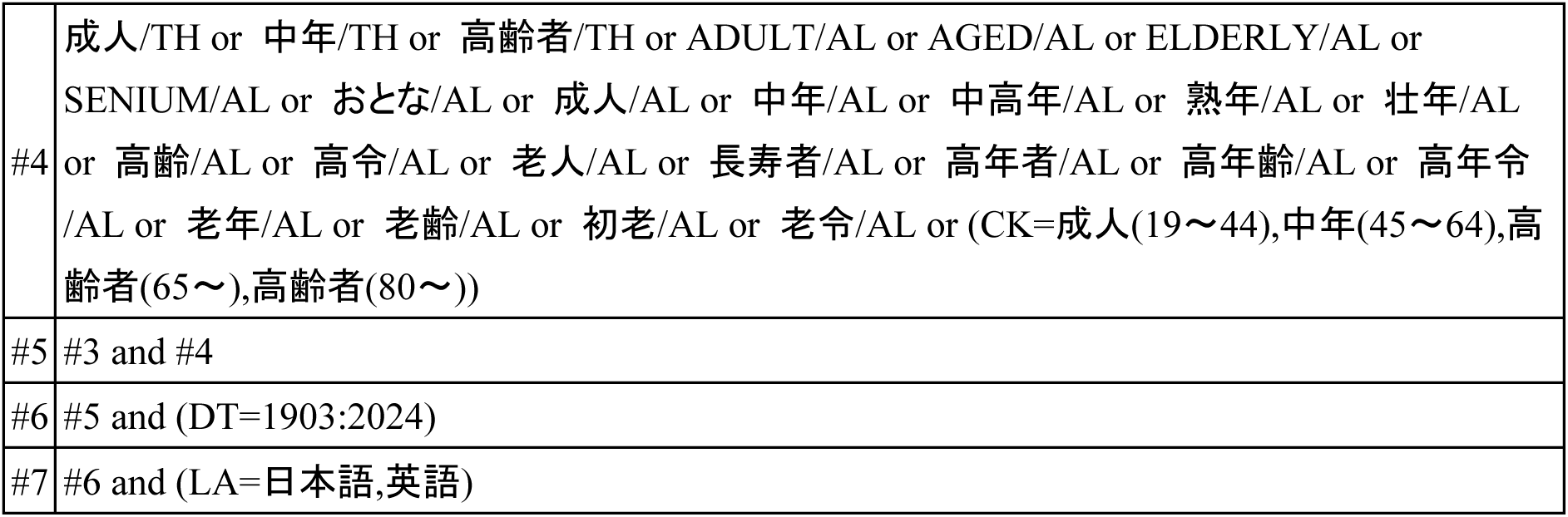
Search strategies.

### Study Selection

Two independent reviewers screened titles and abstracts using Rayyan (https://www.rayyan.ai), followed by a full-text review against prespecified eligibility criteria. Disagreements were resolved through discussion. To ensure methodological independence across KQs, reviewer assignments were managed by the designated Literature Review Team. Eligibility criteria were defined separately for each KQ (Table 2). Eligible study designs included randomized controlled trials (RCTs), large-scale cohort studies, systematic reviews, and meta-analyses, depending on the specific question. For KQ1 and KQ3, studies evaluating the effectiveness of screening or revascularization in asymptomatic adults were included. For KQ2 and KQ4, studies were required to report on harms associated with screening, confirmatory testing, or surgical interventions (e.g., carotid endarterectomy (CEA), carotid artery stenting (CAS)). All studies had to enroll asymptomatic adults without a history of stroke, transient ischemic attack (TIA), or other neurologic symptoms referable to the carotid arteries. Studies limited to symptomatic populations, diagnostic accuracy of screening modalities, or cost-effectiveness were excluded.

**Table 2.**
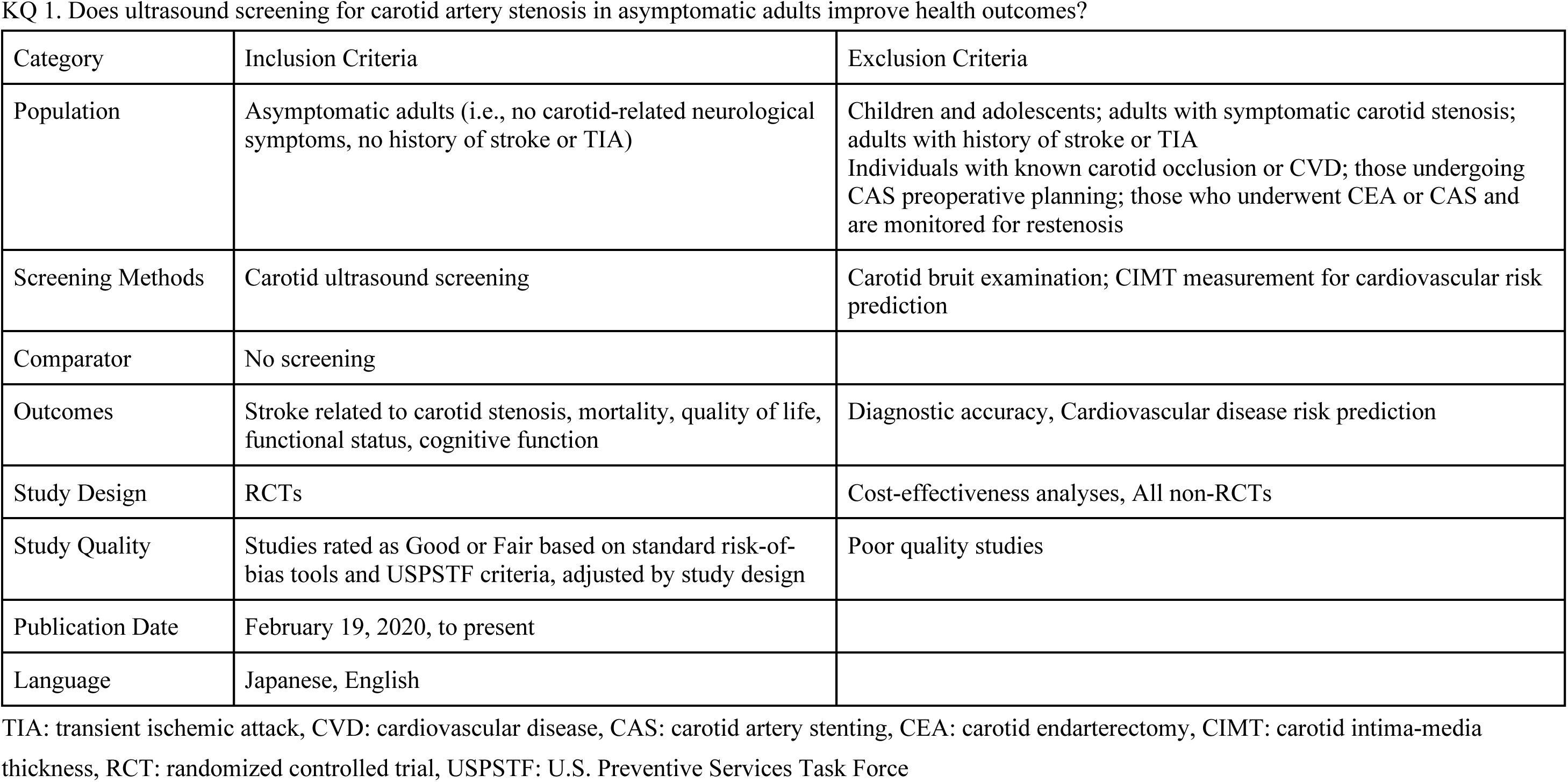
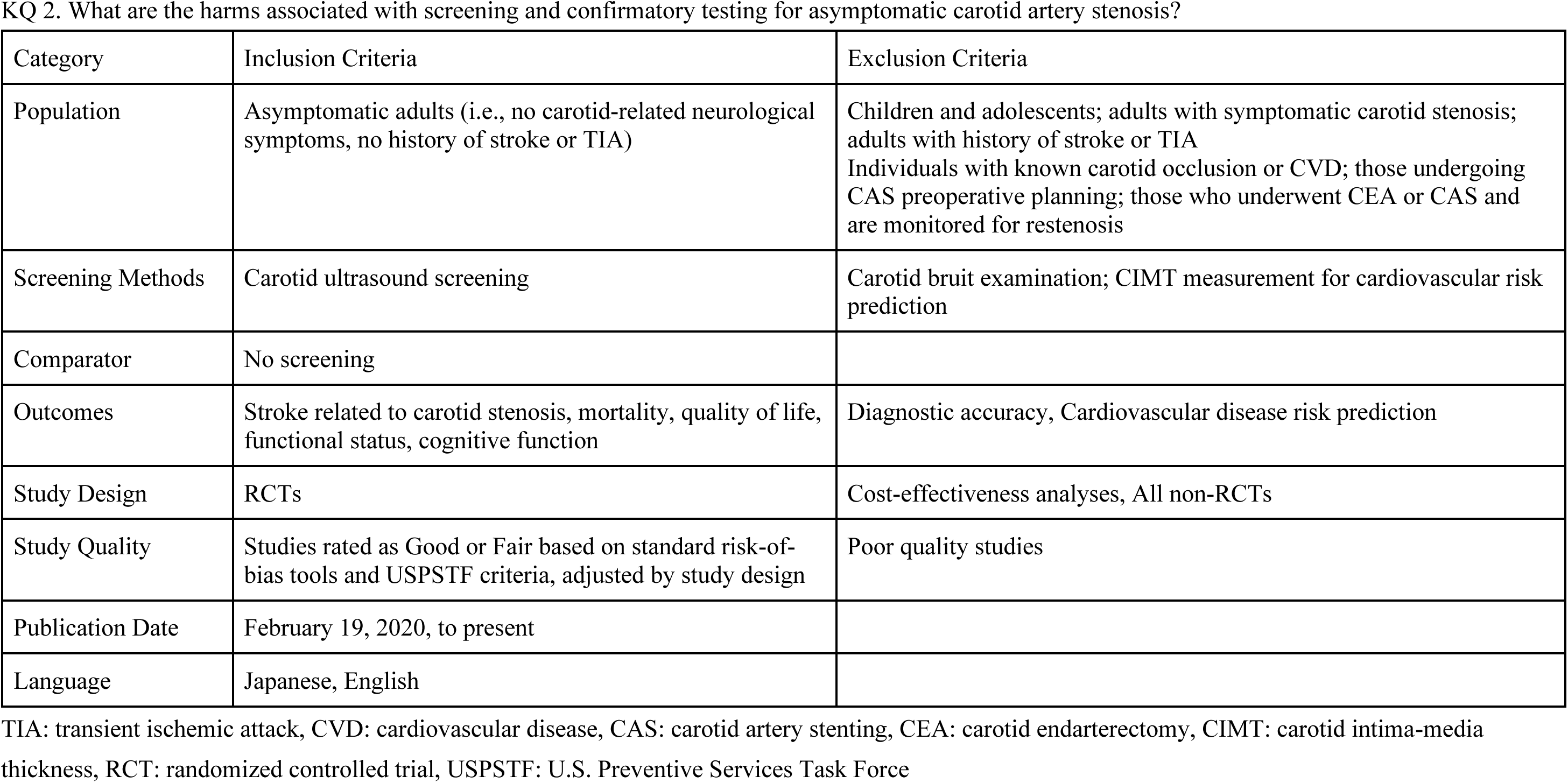
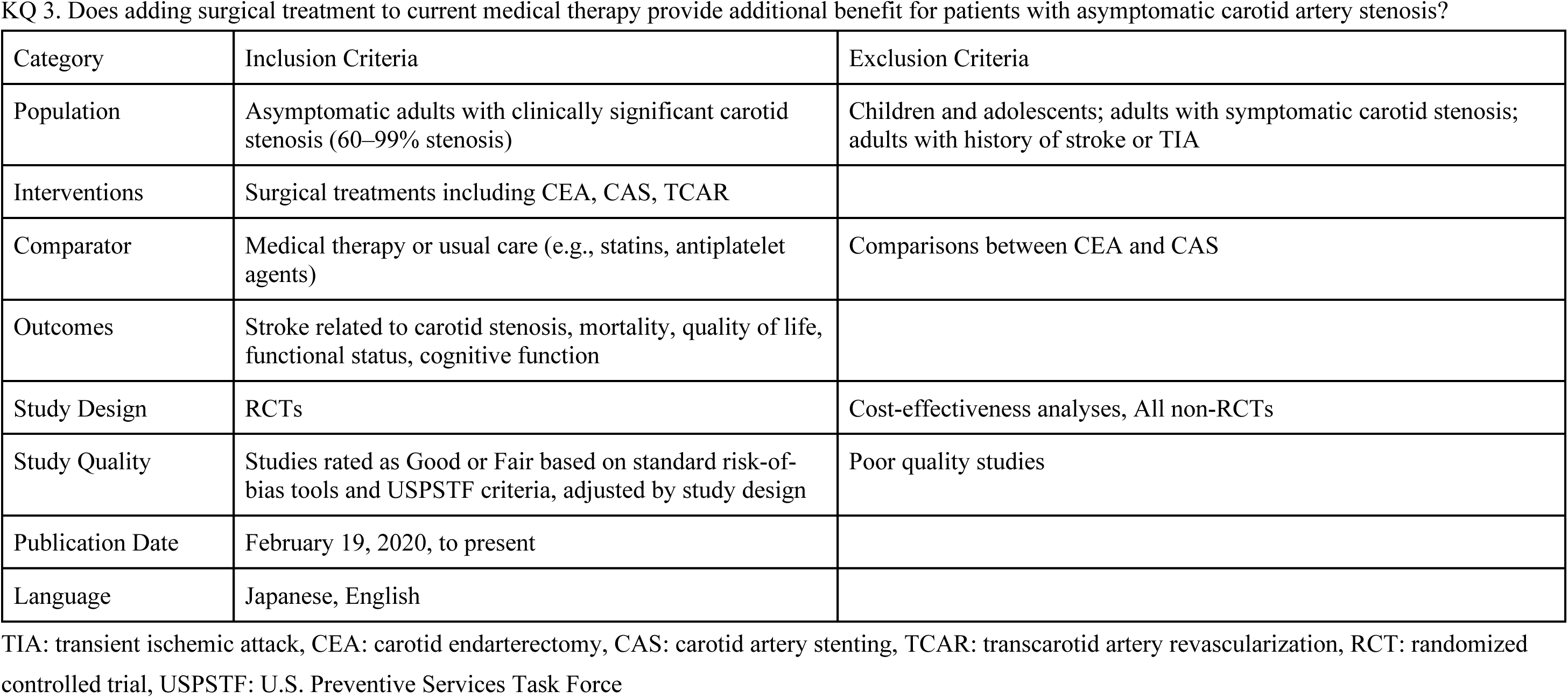
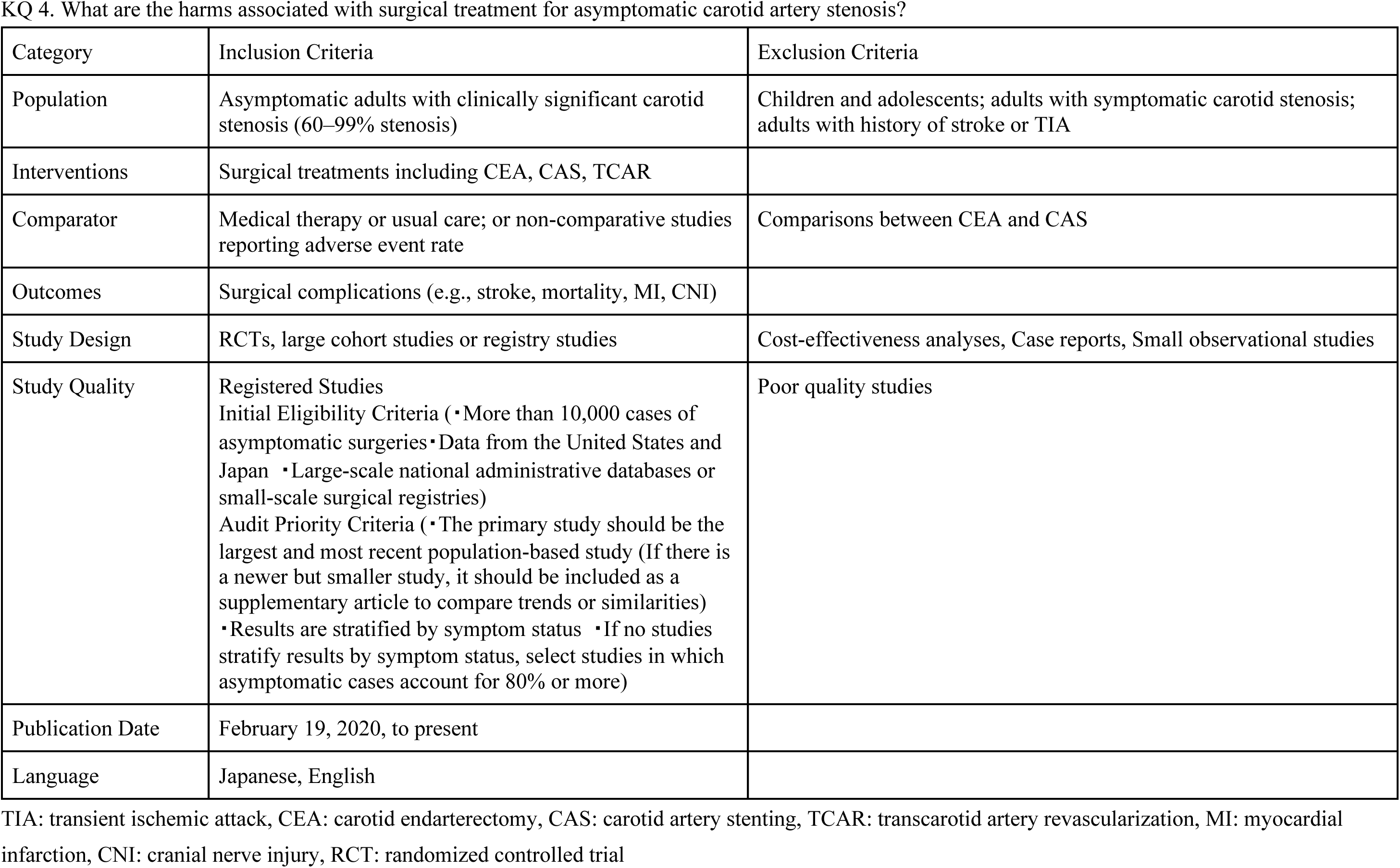
Eligibility criteria were defined separately for each Key Question (KQ)

### Data Extraction and Quality Assessment

Using a standardized form, two reviewers independently extracted relevant data, including study design, population characteristics, intervention, comparator, outcomes, and setting. The risk of bias for each study was assessed independently by two reviewers using design-appropriate tools. The Cochrane Risk of Bias 2 (RoB 2) tool was used for RCTs, and the Risk of Bias in Non-randomized Studies of Interventions (ROBINS-I) tool was applied to non-randomized intervention studies. For large cohort or registry-based studies, a modified Newcastle-Ottawa Scale was used. All disagreements in data extraction or quality assessment were resolved by consensus. Risk of bias was assessed using three tools: RoB 2 categorized studies as having “Low,” “Some concerns,” or “High” risk of bias; ROBINS-I used four categories—“Low,” “Moderate,” “Serious,” and “Critical”; and the Newcastle-Ottawa Scale classified study quality as “Poor,” “Fair,” or “Good.”

### Data Synthesis and Analysis

Findings from newly identified studies were qualitatively synthesized alongside the evidence from the 2021 USPSTF review ^5^. A meta-analysis was not performed in this review, as a relatively recent systematic review conducted by the United States Preventive Services Task Force (USPSTF) was available, and it comprehensively addressed the key outcomes relevant to our review. Additionally, due to the need to complete the review within a limited timeframe for policy relevance, we prioritized a narrative synthesis. For each KQ, results were summarized in descriptive text and tables, focusing on the direction and consistency of findings, study design, and methodological quality.

## Results

### Summary of Evidence Search and Selection

The literature search yielded 764 records. After the removal of duplicates, 666 titles and abstracts were screened. (PRISMA flow diagrams for each KQ are available in Figure 2). No studies met the criteria for full-text review for KQ1 (benefits of screening) or KQ2 (harms of screening). For KQ3 (benefits of revascularization) and KQ4 (harms of revascularization), 6 and 27 studies underwent full-text review, respectively. Ultimately, 1 RCT ^6^ and 5 observational studies ^7–11^ were included in the final synthesis. A detailed list of included studies and their findings is provided in Table 3.

**Figure 2.**
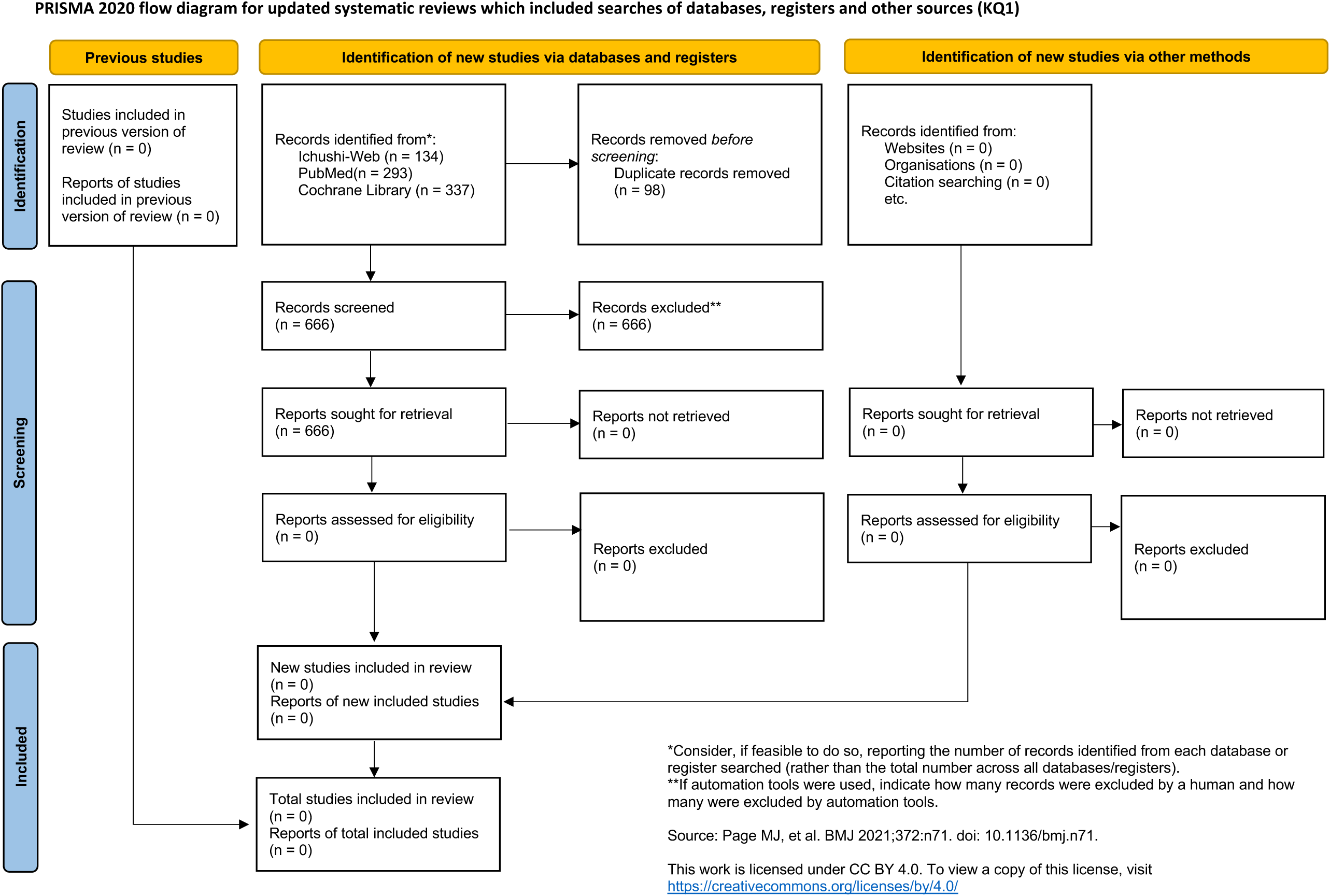
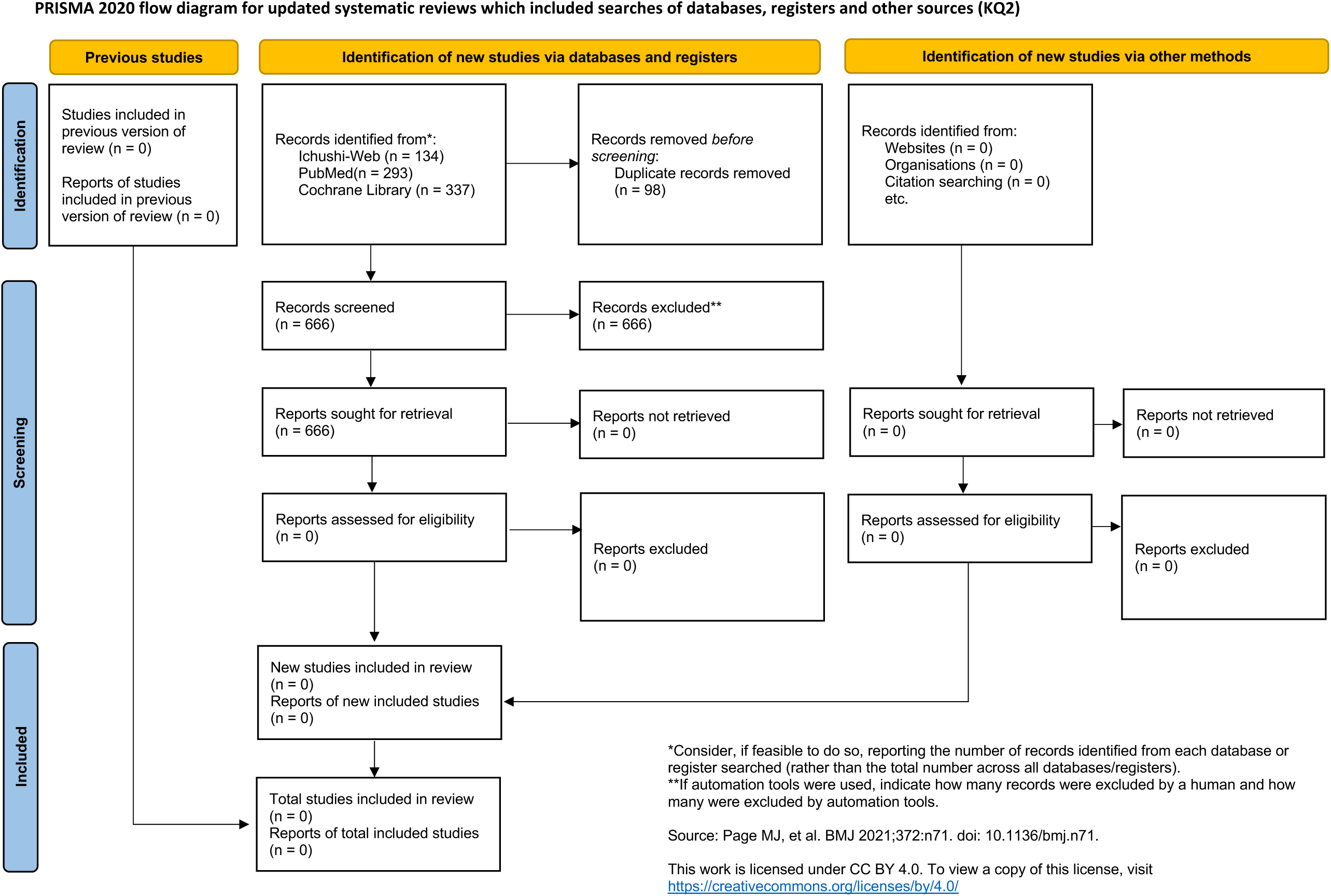
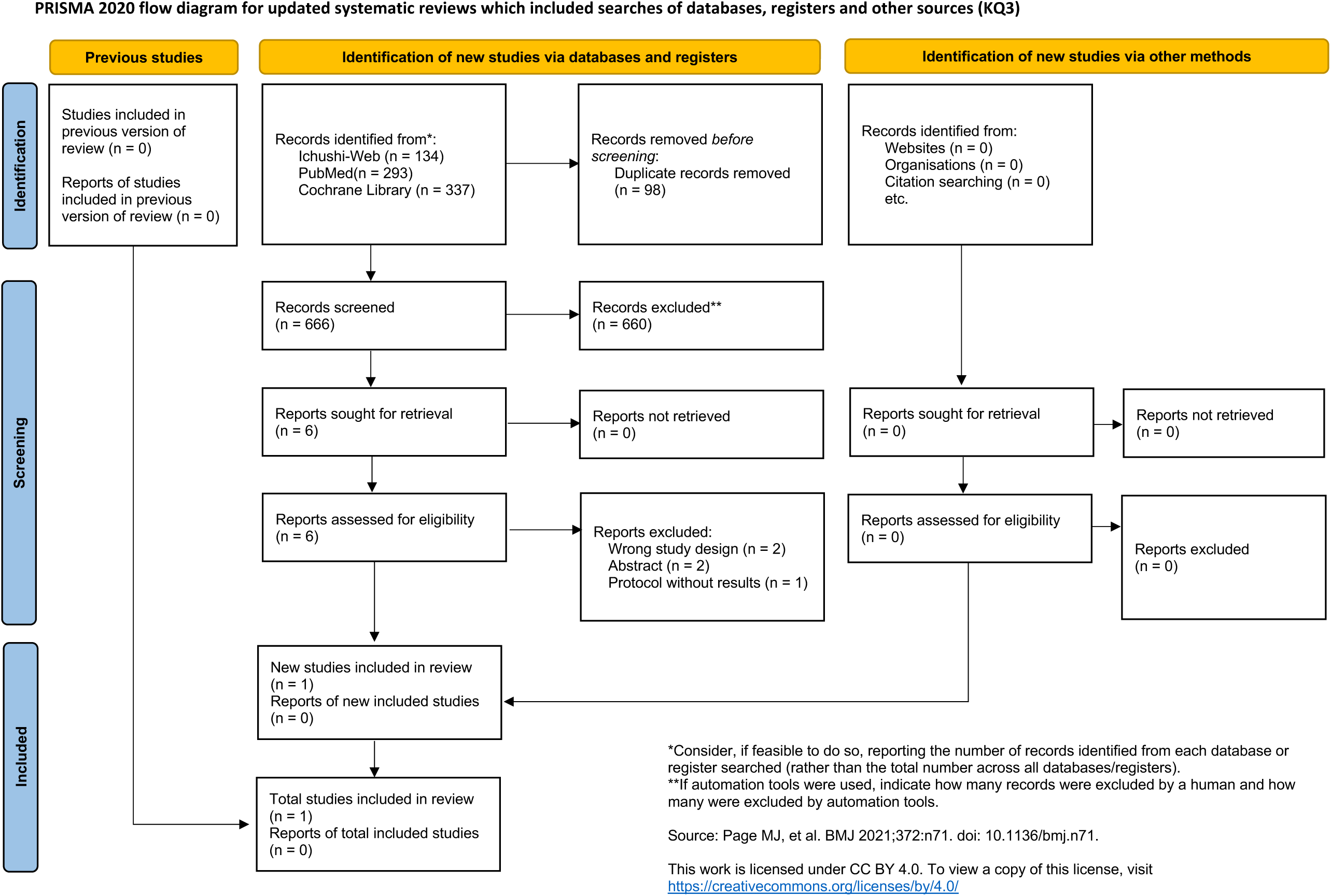
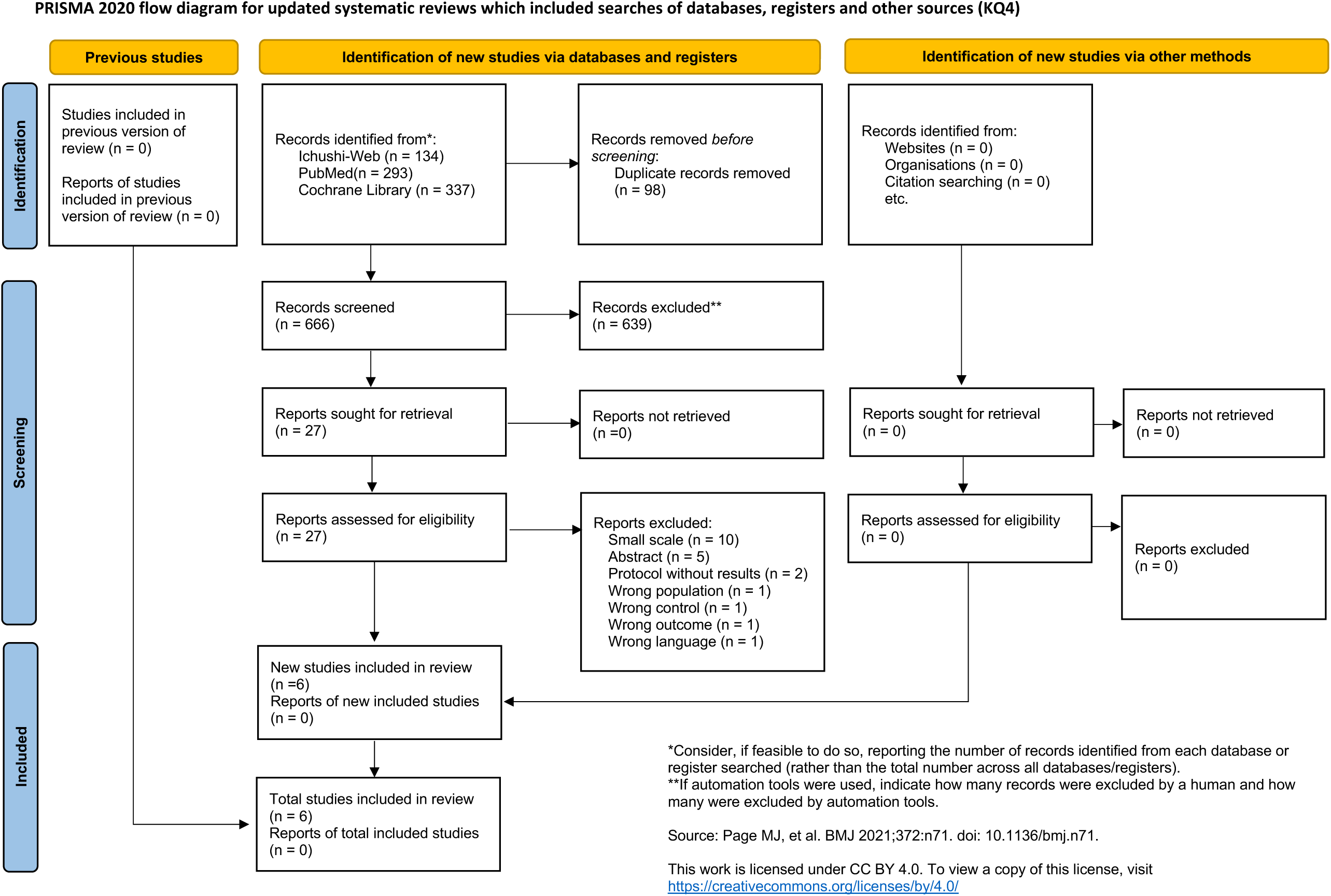
PRISMA flow diagrams for each Key Question (KQ)

**Table 3.** Detailed list of included studies.

| Study | Study design | Country | Type of Surgical Procedure | Follow-up Duration | Setting and Study Population | No. of ACAS Participants | Primary Outcome(s) | RoB | COI Disclosure |
| --- | --- | --- | --- | --- | --- | --- | --- | --- | --- |
| Reiff,2022 | RCT | Austria<br>Germany<br>Switzerland | CEA<br>CAS | 5years | ACAS of 50–70% or more | 513 | Stroke or cerebral infarction within 30 days<br>Ipsilateral stroke within 5 years | High | Disclosures reported<br>Funding reported |
| Keyhani, 2020 | Cohort | USA | CEA | 5years | ACAS ($\geq 70\%$) from the US Department of VA and Medicare data | 5221 | Ipsilateral stroke | Fair | No disclosures reported |
| Poorthuis, 2020 | Cohort | USA<br>Canada<br>Europe | CEA | 30days | Individual patient data from four RCTs targeting ACAS $\geq 50\%$ | 3694 | Stroke and death | Fair | Disclosures reported<br>Funding reported |
| Goicoechea, 2022 | Cohort | USA | CEA<br>CAS | 30days | ACS NSQIP* | 100637 | Stroke | Good | Disclosures reported |
| Deery, 2021 | Cohort | USA | TCAR | 30days | Carotid artery stenosis $\geq 70\%$ in the US VQI | 8460 | Stroke, death, and myocardial infarction within 30 days | Good | Disclosures reported |
| Chang, 2023 | Cohort | USA | CEA | 5years | ACAS $\geq 70\%$ in KPNC | 3824 | Stroke | Good | Disclosures reported |

ACAS: asymptomatic carotid artery stenosis, RoB: risk of bias, COI: conflict of interest disclosure, RCT: randomized controlled trial, CEA: carotid endarterectomy, CAS: carotid artery stenting, VA: Veterans Affairs, ACS NSQIP: American College of Surgeons National Surgical Quality Improvement Program, VQI: Vascular Quality Initiative, KPNC: Kaiser Permanente Northern California, \* Degree of stenosis not reported.

### Results of Included Studies

#### Benefits of Screening

Key Question 1. Does ultrasound screening for carotid artery stenosis in asymptomatic adults improve health outcomes?

No RCTs evaluating the benefits of screening for ACAS were identified within the protocol-defined search period.

#### Harms of Screening

Key Question 2. What are the harms associated with screening and confirmatory testing for asymptomatic carotid artery stenosis?

No RCTs assessing the harms of screening for ACAS were identified within the protocol-defined search period.

#### Incremental Benefits of Revascularization

Key Question 3. Does adding surgical treatment to current medical therapy provide additional benefit for patients with asymptomatic carotid artery stenosis?

A single RCT was identified that assessed the incremental benefits of revascularization for ACAS ^6^. The SPACE-2 trial, a 3-arm parallel-group study, enrolled 513 participants comparing CEA plus best medical therapy (BMT), CAS plus BMT, and BMT alone. The study reported on outcomes including stroke, mortality, and functional status but did not assess quality of life or cognitive function. The cumulative 5-year incidence of stroke or death within 30 days was 2.5% in the CEA+BMT group, 4.4% in the CAS+BMT group, and 3.1% in the BMT-alone group; these differences were not statistically significant. When considering ipsilateral ischemic stroke alone, the 5-year cumulative incidence was 2.0% for CEA plus BMT, 4.4% for CAS plus BMT, and 3.1% for BMT alone. Although the incidence was slightly lower in the CEA group, the differences were not statistically significant. Similarly, there were no statistically significant differences in all-cause mortality (7.6% for CEA+BMT, 9.3% for CAS+BMT, and 8.0% for BMT alone) or the cumulative incidence of stroke with a modified Rankin Scale score >2 at 30 days (1.0%, 1.7%, and 2.0%, respectively). This study was judged to be at a high risk of bias (Figure 3).

**Figure 3.**
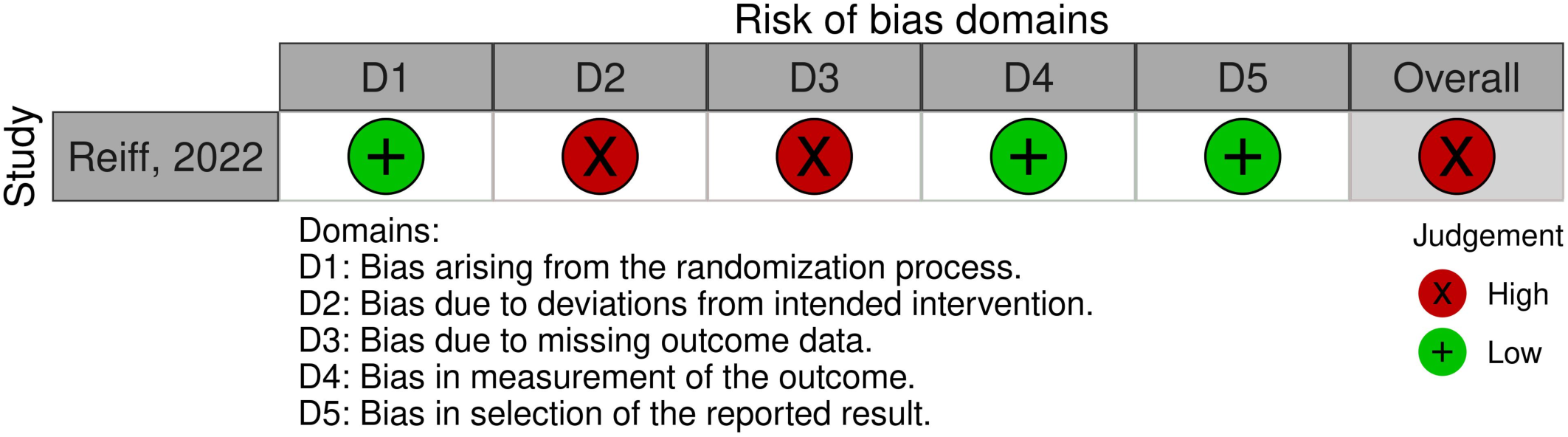
Risk of bias for the randomized clinical trial.

#### Harms of Revascularization

Key Question 4. What are the harms associated with surgical treatment for asymptomatic carotid artery stenosis?

One RCT ^6^ (also included in KQ3) and 5 observational studies ^7–11^ were identified.

Stroke or Death Within 30 Days: Three studies reported this composite outcome. The RCT by Reiff et al. ^6^ found no statistically significant difference between CEA or CAS compared to BMT. Two large retrospective cohort studies reported incidences of 2.5% ^10^ and 2.8% ^11^, respectively, though neither included a non-surgical comparator group.

Stroke or Death Within 1 Year: One large registry-based study (n=8,460) reported a 1.2% incidence of this outcome among patients undergoing transcarotid artery revascularization (TCAR) ^8^.

Stroke (Perioperative or Long-Term): Six studies reported on stroke as a harm. The RCT showed no statistically significant difference between the intervention and control groups ^6^. Incidence rates in the observational studies ranged from 0.9% to 7.5% ^8–11^.

Death: Five studies addressed this outcome. The RCT found no statistically significant differences ^6^. Mortality rates in the observational studies were 0.3% ^8^, 0.7% ^9^, and 0.9% ^11^.

Myocardial Infarction (MI): Four studies reported MI rates. The RCT ^6^ found no statistically significant differences, and 3 observational studies reported rates from 0.3% to 0.9% ^8, 9, 11^.

Cranial Nerve Injury: One study ^8^ reported a 0.1% incidence of cranial nerve injury following TCAR.

Other outcomes evaluated in the randomized controlled trial by Reiff et al. ^6^ included ipsilateral stroke, ischemic stroke, disabling stroke, ipsilateral disabling stroke, ischemic stroke or vascular death, restenosis and progression of stenosis, vascular death, transient ischemic attack (TIA), and ipsilateral TIA. For all of these outcomes, no statistically significant differences were observed in either relative or absolute risk between CEA and CAS.

The risk of bias assessments for the 5 observational studies are summarized in Table 4.

**Table 4.** Risk of bias assessments for the five observational studies.

|  | Representativeness<br>of exposed cohort | Selection of non-<br>exposed cohort | Ascertainment<br>of exposure | Comparability of cohorts on the<br>basis of the design or analysis | Assessment<br>of outcome | Adequacy of<br>follow up | Score and levels<br>of evidence |
| --- | --- | --- | --- | --- | --- | --- | --- |
| Keyhani, 2020 | ★ | 0 | ★ | ★★ | ★ | ★ | 6, Fair |
| Poorthuis, 2020 | ★ | 0 | ★ | 0 | ★ | ★ | 4, Fair |
| Goicoechea, 2022 | ★★ | ★ | ★ | 0 | ★ | ★★ | 7, Good |
| Deery, 2021 | ★★ | ★ | ★ | ★ | ★ | ★★ | 8, Good |
| Chang, 2023 | ★★ | ★ | ★ | ★★ | ★ | ★★ | 9, Good |

## Discussion

### Summary of Findings

In 2021, the USPSTF recommended against screening for ACAS in the general population (Grade D), concluding that the potential benefits do not outweigh the harms ^5^. This review aimed to provide an up-to-date and objective evaluation of the current evidence on screening and management of asymptomatic carotid artery stenosis (ACAS) in Japanese primary care, with a focus on internal validity. However, no eligible studies were identified that directly assessed the benefits or harms of ACAS screening. The available evidence on treatment is also limited. The single RCT that assessed the incremental benefit of revascularization (SPACE-2) was terminated prematurely and possessed substantial methodological limitations, including a high risk of bias ^6^. In five registry-or claims-based observational studies ^7–11^, the 30-day incidence of stroke or death following revascularization was 2.5% to 2.8%. Perioperative stroke, death, and myocardial infarction occurred in 0.9% to 2.3%, 0.3% to 0.9%, and 0.3% to 0.9% of patients, respectively. These findings were consistent with the conclusions of the 2021 USPSTF review.

The internal validity of the studies included in this systematic review was limited by variations in risk estimation and outcome assessment methods across studies, potentially affecting the reliability of their findings. The RCT by Reiff et al. ^6^ suffered from a small sample size and high attrition, undermining its statistical power and introducing potential selection bias. Furthermore, it was judged to be at high risk of bias due to deviations from the intended intervention and issues with missing outcome data, raising concerns about the reliability of its findings. The wide confidence intervals for both relative and absolute risks indicate substantial uncertainty. Among the observational studies, several lacked appropriate comparator groups and had inadequate adjustment for confounding, limiting causal inference ^10, 11^. The study by Poorthuis et al. ^11^ did not adjust for any prognostic variables. In contrast, studies by Goicoechea et al. ^9^, Deery et al. ^8^, and Chang et al. ^7^ employed more rigorous methods for confounding adjustment and were thus considered to have higher internal validity. Regarding the reliability of outcome assessment, RCT were subject to risks such as lack of blinding, while retrospective studies often had limitations in consistency and objectivity of outcome evaluation. In particular, rare outcomes such as surgery-related complications and cranial nerve injury may be prone to reporting bias. Furthermore, the Vascular Quality Initiative (VQI) database used in the study by Deery et al. was based on clinical data from selected hospitals, physicians, and cases.

Overall, the internal validity of the studies included in this review was subject to several limitations, including small sample sizes and high attrition rates in RCTs, as well as sample bias, absence of comparison groups, and inadequate adjustment for confounding in retrospective observational studies.

### Limitations

This review has several important limitations. First, the absence of eligible published studies involving Japanese populations precludes a direct assessment of the effectiveness or harm of ACAS screening in Japan. The review relied on studies conducted primarily in Western countries, whose populations may differ from Japan’s in terms of genetic background, healthcare systems, and risk factor profiles. While one Japanese study assessing the safety of CAS in elderly patients ^12^ was excluded due to small sample size, its finding of a 1.85% incidence of major stroke, myocardial infarction, or death within 30 days post-CAS aligns with the range reported in this review. In addition, although the literature search in Ichushi-Web did not impose a publication date restriction, studies published before February 19, 2020, were excluded at the full-text screening stage based on the protocol-defined eligibility criteria. One such study was the Japan Carotid Atherosclerosis Study (JCAS) ^13^, a large multicenter prospective registry conducted in Japan. Although JCAS formally met the threshold for a “fair” quality rating under the modified Newcastle-Ottawa Scale, the absence of any adjustment for confounding variables (0 points for comparability), along with the potential for selection bias, raised concerns regarding its internal validity. Including this study in the evidence synthesis would not have altered the overall conclusions of the review.

Second, this review focused on procedural harms (e.g., stroke, death), but did not systematically assess non-physical harms such as the psychological burden of a new diagnosis, overdiagnosis, or the resource utilization associated with screening programs.

Third, the included studies often had long data collection periods. Continuous advancements in both medical therapy (e.g., high-intensity statins) and surgical techniques may have lowered event rates in contemporary practice, potentially altering the risk-benefit from what is reported in the literature.

### Conclusions

There is a lack of direct evidence evaluating the effectiveness and harms of screening for asymptomatic carotid artery stenosis. Furthermore, the evidence regarding the benefits and harms of adding revascularization to optimal medical therapy is limited by early termination of trials and methodological limitations, thereby reducing the internal validity and warranting cautious interpretation of the findings.

## Disclaimer

This review was conducted independently of any funding organization. The findings and conclusions expressed herein do not represent the official positions of the authors’ affiliated institutions.

## Type of contribution of the authors

Study Concept and Design: All authors

Acquisition, Analysis, or Interpretation of Data: All authors

Drafting of the Manuscript: Yusuke Saishoji

Critical Revision of the Manuscript for Important Intellectual Content: Tomoharu Suzuki, Saori Nonaka, Shunsuke Suya, Takashi Kitagawa Study Supervision: Yusuke Saishoji

Dr. Saishoji had full access to all study data and takes full responsibility for the integrity of the data and the accuracy of the data analysis.

IRB Approval Code and Name of the Institution

Ethical review was deemed unnecessary by The Ethical Committee of Kurume University (health care & medical ethics), as this work is based solely on a literature review and does not involve human subjects or identifiable personal data.

## Acknowledgments

This review was conducted as part of the JPPSTF Project, which was commissioned by EVIDENCE STUDIO, a general incorporated association whose purposes include optimizing public healthcare expenditures in Japan. The contract was formally established with Kurume University, with which Kei Mukohara, the chair of the JPPSTF, is affiliated.

## Use of AI-assisted tools

During the preparation of this manuscript, an AI-assisted language model (ChatGPT, OpenAI, accessed in 2025) was used to assist in editing and improving the clarity and structure of the text. All content was critically reviewed and finalized by the authors, who take full responsibility for the accuracy and integrity of the manuscript.

